# Microvascular dysfunction induces a hyperdynamic circulation; a mathematical exploration

**DOI:** 10.1101/2024.07.22.24310841

**Authors:** Ivor Popovich

## Abstract

**Background:** The discordance between the macrocirculation and microcirculation in septic shock has been recognised but never explained. I present a novel mathematical hypothesis as to how heterogenous microcirculatory flow distribution directly induces a hyperdynamic circulation and how elevated central venous pressure induces microcirculatory dysfunction.

**Methods:** I explore the tube law and modified Poiseuille resistance for compliant blood vessels. Using these equations a new equation is developed incorporating time constants, elastance of the vessel, unstressed volume and wave reflections that demonstrates the relationship between volume of a microcirculatory vessel and total flow through it.

**Results:** The relationship is demonstrated to be constant at zero until the unstressed volume is reached after which it increases exponentially. By considering n of these vessels in parallel, I demonstrate that the summed flow is minimised when flow is equally distributed among the n vessels, while it is maximised when all flow goes through one vessel alone, thereby demonstrating that heterogenous microvascular perfusion leads to increased total flow. It is shown that if conditions of wave reflection are right then a hyperdynamic circulation with high cardiac output develops. It is also demonstrated that high central venous pressure increases wave reflections and necessarily leads to microvascular perfusion heterogeneity if cardiac output is to be maintained.

**Conclusions:** Microvascular impairment in septic shock directly leads to a hyperdynamic circulation with high cardiac output. High central venous pressures impair the microcirculation. Decades of clinical findings can now be explained mathematically. Implications for hemodynamic therapy for septic shock are discussed.

**Clinical Perspective:** Research in septic shock has focussed on two main components of the circulation; the macrocirculation and microcirculation. Very discordant findings have been observed between the two, to the point that the term ‘uncoupling’ has appeared in the literature in reference to these two circulations. Thus far nobody has put forward a satisfactory physiological explanation as to the mechanism of this discordance. There is a need to understand the physiological mechanisms to guide future attempts and research into methods of resuscitating the septic patient in order to improve circulatory function.

This work provides the first theoretical mathematical groundwork as to not only how these circulations are linked, but how they directly influence one another. This sets the framework for future clinical and basic science research and helps us understand how our current resuscitation strategies may work to restore the microcirculation, and when they may start to impair it.

## Introduction

The aim of resuscitating the shocked patient is to restore cellular oxygen delivery and energy utilisation by restoring cellular blood flow (1). The adequacy of the circulation in this regard can be assessed using either ‘macrocirculatory’ or ‘microcirculatory’ variables. The former consists of things easily measured in clinical practice that reflect global oxygen delivery i.e. heart rate, cardiac output, blood pressure, and central venous pressure. The latter is difficult to measure at the bedside and includes parameters only used in research studies, such as the functional capillary density and perfused vessel density (2). While assessment of the microcirculation is the more important of the two, as it most closely reflects the end points of resuscitation, it is generally believed that optimising macrocirculatory parameters will optimise the microcirculation.

Increasing evidence has accumulated that this is not the case. It is now well-known that in septic shock without concurrent hypovolemia that a hyperdynamic (high cardiac output) circulation develops, however severe microcirculatory derangements persist and are directly associated with mortality. These consist of reduced vessel density and heterogeneous flow distribution. Organ dysfunction develops despite elevated organ blood flows (3-6). The nature of these paradoxical findings has never been fully explained. Traditional resuscitation paradigms have focussed on fluid resuscitation to increase central venous pressure (CVP) in order to further increase cardiac output and organ perfusion. Recently, it has been shown in sepsis that a high central venous pressure, independently of other hemodynamic variables, impairs organ function, particularly renal function (7). This adds to identical findings in another group of patients, those with chronic heart failure, where the association is also independent of cardiac output (8).

The aim of this study is to put forward a mathematical explanation as to how the macrocirculation and microcirculation are linked and to explain why elevated central venous pressures may impair the microcirculation despite being associated with improved global hemodynamics.

## Methods *(Mathematical background)*

Understanding the novel mathematics in this paper requires us to explain some existing concepts. The circulation is composed of travelling flow (Q) and pressure (P) waves. At any given anatomical location in the circulation the relationship between the magnitude and shape of the pressure and flow waves is determined by the impedance (Z). The equation relating the two is;

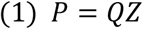

P and Q are wave functions and Z is a complex number (i.e. it contains both real and imaginary parts) and multiplying Q by Z is not a straightforward exercise. However we can simplify this greatly by considering only the mean pressure and flow, which is what we are interested in, rather than the whole wave. In doing this Z reduces to a real function which is easy to multiply.

A problem is encountered however as we pass along the circulation; the impedance changes. Imagine a junction between 2 segments of the circulation. Impedance of the upstream segment is Z_1_ and the impedance of the downstream segment is Z_2_. At the junction the flow and pressure are the same yet there are two different values of Z. Mother nature solves this paradox by reflecting some of the upstream waves back at the junction (9). Thereby each segment of the circulation contains both a forward and backward travelling wave, and therefore a forward and reverse mean flow. The net flow is the same in each segment and results from subtracting the reverse flow from the forward flow.

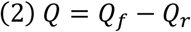

Where Q is the net flow, Q_f_ is the forward flow and Q_r_ is the reverse flow.

In contrast the forward and reverse pressures are additive;

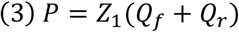

This allows each segment to maintain its own Z, while allowing the net flow and net pressure at the junction to be equal between them. For example, the upstream segment may have a forward flow of 5 L/min and a reverse flow of 2 L/min, while the downstream segment may have a single forward flow of 3 L/min. In both cases the net flow is the same. In fact the size of the reverse flow relative to the forward flow (C), assuming that the downstream segment does not have any reflected waves of its own, can be defined as follows;

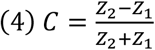

The mathematical derivation of this is straightforward (10) and not reproduced here. The downstream segment in fact does have reflected waves of its own (from a segment even further downstream from itself) so the equation is not quite so simple. However the same principle remains; the greater the impedance of a downstream segment compared to the impedance of the upstream segment, the stronger the reflected waves. This is the same principle that governs the clinical utility of ultrasound waves.

Now we must turn our attention to understanding the nature of the compliant segments of the circulation. The circulation is simplified for the benefit of clinicians’ understanding into a linear model. Linearity means that the flow through a vessel does not affect the characteristics of the vessel. Taking a stiff artery, and assuming that the vascular tone of the artery remains constant, doubling the flow through said artery will not change the volume of blood in the artery, its compliance, nor its resistance. A doubling of flow leads to a doubling of the pressure gradient. However in the microcirculation, composed of arterioles, capillaries and venules, this is not the case (11). Haemodynamics here are highly non-linear. Because the microcirculation (in particular the venules) is highly compliant, increasing flow through it will increase its volume, which will decrease its resistance, and the resultant change in the pressure gradient is not easily predictable.

The importance of these observations has already been recognised in the venous function work of Guyton and other authors who have followed on from this (12). The venous system is recognised as having an unstressed volume, which does not generate any pressure in the veins. Anything above this volume is the stressed volume, which begins to generate pressure and therefore venous return. Increasing venous return increases the preload available to the heart, which will respond by increasing its cardiac output (in the long run venous return and cardiac output are the same) (13). The stressed volume can either be increased by fluid or blood infusion, or by catecholamine induced increases in venous tone, the latter decreasing the venous compliance and the unstressed volume.

## Results *(Mathematical development)*

The pressure-volume relationship of veins has been extensively modelled in the works of Pedley et al. (14-15). The same principle can be applied to the other compliant/non-linear vessels of the microcirculation. Pressure as a function of volume of a vessel segment is described with the following equation;

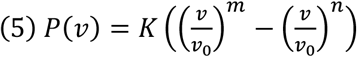

This is known as the tube law.

P is the transmural pressure, although we assume the external pressure to be zero, so the internal pressure equals the transmural pressure. K is a constant, v is the volume of the vessel segment, v_0_ is the unstressed volume, m is some positive integer and n is some negative integer.

In a non-compliant tube the resistance is described by the traditional Poiseuille equation. This is inadequate for modelling the resistance in compliant vessels. As the volume and/or transmural pressure of a compliant vessel decreases, the vessel undergoes partial collapse. The shape of the cross-sectional area changes from tubular to elliptical, which imparts greater viscous resistance to the tube. Thankfully Pedley has derived a modified Poiseuille equation to account for this behaviour (14). Resistance as a function of volume is described by;

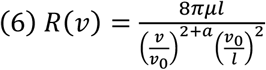

R is the resistance of the vessel segment, µ is the viscosity, l is the length of the segment (assumed to remain constant), and a is some positive integer (0.5 was the value used by Pedley et al. (14)).

There are other assumptions inherent in using the modified Poiseuille equation to model resistance. Firstly flow must be laminar and secondly the fluid involved must be Newtonian. Blood is known to be non-Newtonian, meaning that its viscosity falls as its shear stress increases. Turbulent blood flow also occurs in parts of the circulation, although in the microcirculation Reynolds numbers have been found to be in the range indicating laminar flow (16). Either way these assumptions produce only minor deviations from observed values and are acceptable in modelling the circulation (17).

All distensible tubes with both a compliance and a resistance that either empty or fill with flow will exhibit a time constant τ. Such a tube will empty at an initial rate that then decreases in an exponential fashion with time, until the tube has completely emptied. The time constant represents the time that would be required for the system to empty to zero if emptying had continued at that initial rate (18). Therefore the initial flow rate of emptying in units of volume per time is;

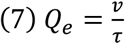

Q_e_ is the initial flow rate of emptying and v is the initial volume.

Usefully, the time constant is determined by the elastance and resistance of the tube.

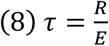

E is elastance. The elastance of a distensible tube segment is simply the derivative of its pressure with respect to volume. Therefore;

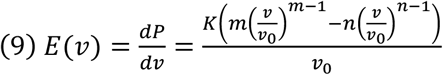

Eq. 9 is simply the first derivative of Eq. 5.

When the circulatory system is in steady state the mean volume of a compliant tube segment is constant. Therefore the rate of emptying of the tube segment equals the inflow into the segment.

Since the segment is not changing volume, the emptying rate continues at the initial rate. We know the elastance and resistance of the tube segment, therefore;

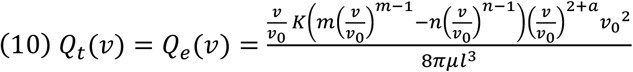

Q_t_ is both the inflow and outflow and therefore the total flow through the segment. This is where we need to recall the forward and reverse wave phenomena covered in the previous section. The tube segment both empties and fills in both the forward and reverse directions. Therefore;

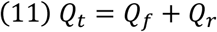

If we plot Eq. 10 we observe the following (Fig 1.).

**Figure 1.**
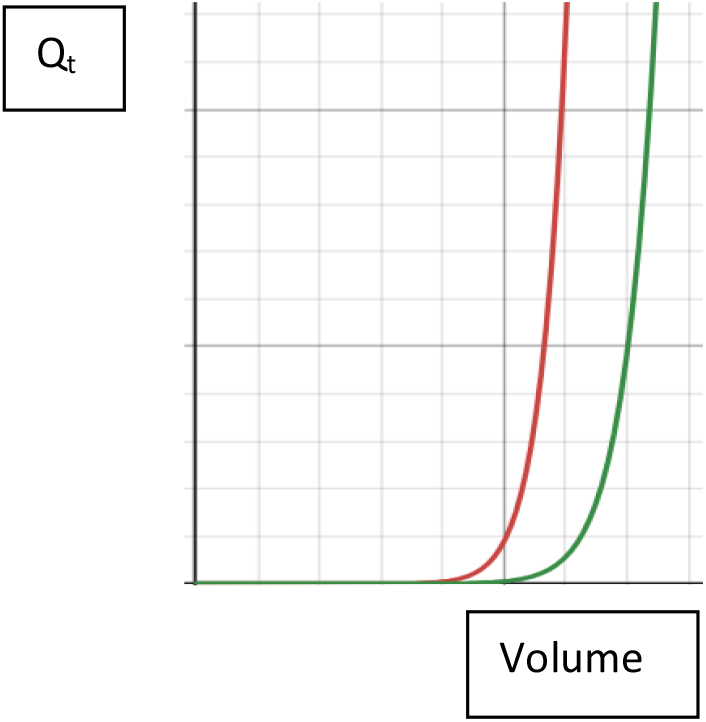
Flow vs volume of a compliant tube segment.

We see that as the tube fills with volume this does not generate any flow at first until it reaches a critical volume, after which flow starts to rise exponentially. By choosing an appropriately small value for K in the equation, this critical volume becomes v_0_. The difference between the red curve and the green curve is that v_0_ has been decreased in the red curve.

That Q_t_ is the summation of both the forward and reverse mean flow may be difficult for some readers to accept. We can see however how it is consistent with our clinical observations. If the generated flow was only the net forward flow then we would expect an increase in volume of compliant vessels to always result in an increased cardiac output. However in a failing heart a large volume may exist in the venous circulation despite a low cardiac output; in this case the reverse flow is large, so Q_t_ remains high, however the net forward flow is low.

Now let us consider the vascular bed of a single organ. Considering the microvascular, compliant portion, we have a bed made up of ŋ vessel segments in parallel, each represented by Eq. 10. Since they exist in the same bed, let us assume they are identical in all the constants in Eq. 10. That is, the only thing that changes between them is their volume. The Q_t_ of that organ bed as a whole will be the summation of Q_t_ in the ŋ vessel segments.

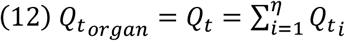

Any organ bed will contain a given total volume, which we will call B, which can be distributed amongst the ŋ vessel segments in any combination. The pattern of volume distribution will therefore affect the total Q_t_. For example, to take an extreme hypothetical, if there are two total vessel segments, the total flow will be different if the total volume is distributed evenly amongst them compared to if all the volume is contained in only one of the vessel beds. In fact the total flow can be described with the following equation;

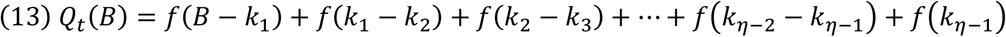

We use the notation *f*() for the simplification of the math that is to follow to represent each *Q*_*ti*_)*v*), or the flow in each vessel segment. The *k* terms are simply a way of splitting up the total volume B amongst the ŋ segments. You need ŋ-1 *k* terms to create ŋ vessel segments, where *B*≥ *k*_1_ ≥ *k*_2_ ≥⋯≥ *k*_*n*−1_.

Now we want to find the values of k that minimise the function Q_t_(B). Taking the partial derivative of the function with respect to any one of the k variables, termed generically as k_ϕ_;

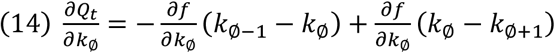

When k_ϕ_ = k_1_, then k_ϕ-1_ = B.

To find the minimum or maximum the partial derivative is set equal to 0. Therefore;

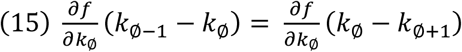

The entire function is either maximised or minimised when all values of k satisfy the above formulation. This occurs when all the terms on the right side of Eq. 13 are identical i.e. when the total volume B is divided evenly among the ŋ vessel segments. Progressing on to the second derivative;

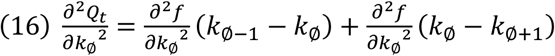

Since *k*_∅−1_ − *k*_∅_ = *k*_∅_ − *k*_∅+1_ at this maximum/minimum, then;

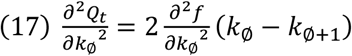

The function *f*(*k*_∅_ − *k*_∅+1_) is just the curve from Fig 1. It can be seen that all order derivatives of this curve will be a similarly shaped curve that also has only positive values. Therefore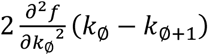 will always be a positive value and *k*_∅−1_ − *k*_∅_ = *k*_∅_ − *k*_∅+1_ must be a local minimum. Total organ Q_t_ is therefore minimised when the total volume, B, of the organ is distributed evenly amongst all ŋ vessel segments.

Plotting the curve graphically for Eq. 13 shows it to be u-shaped. We have identified the value for the minimum. The maximum with respect to any value of k_ϕ_ exists at either end of the curve i.e. when either *k*_∅−1_ − *k*_∅_ = 0 or when *k*_∅_ − *k*_∅+1_ = 0.

If *k*_∅−1_ − *k*_∅_ = 0, then the neighbouring terms to the right become;

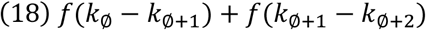

Now to maximise these with respect to *k*_∅+1_ either *k*_∅−1_ − *k*_∅+1_ = 0 or *k*_∅+1_ − *k*_∅+2_ = 0. Let us say now that *k*_∅+1_ − *k*_∅+2_ = 0. The new adjacent pair of terms becomes

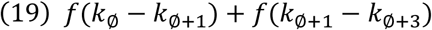

This can again be maximised with respect to *k*_∅+1_

We can continue this exercise in both directions maximising pairs of adjacent *f*() terms with respect to different k terms until we are left with just one term that contains the entire volume B. Therefore total organ Q_t_ is maximised when the total volume, B, of the organ is distributed into just one vessel segment and the other vessel segments contain zero volume.

## Discussion

I have demonstrated a model wherein the total flow in a compliant segment of the microcirculation is dependent on its volume. The total flow represents the mean of both forward and reverse travelling waves. Total flow in an organ bed is related to the distribution of volume, and therefore arterial inflow, amongst parallel microvascular segments, as well as total volume. High total flow results from heterogeneous volume distribution, while low total flow results from homogenous volume distribution. For volume to be redistributed, arterial inflow into the microvasculature must be redistributed. Therefore a better way to think of the overall findings is that microvascular flow heterogeneity increases total flow, and vice-versa. Despite a high overall cardiac output, segments of the microcirculation may receive inadequate flow while others receive excessive flow.

This redistribution phenomenon provides a much needed theoretical model that explains the contradictory and paradoxical associations of the microcirculation and macrocirculation. Clinicians such as myself have long struggled to understand why patients with elevated cardiac output, warm peripheries and increased blood flow to all organ beds nevertheless exhibit clear signs of organ hypoperfusion. Maldistributed flow to the microcirculation has long been recognised, however this is the first work to suggest that there is a causative association; maldistribution directly induces a hyperdynamic circulation.

The traditional explanation for the hyperdynamic circulation has centred on generalised vasodilation causing low systemic vascular resistance and therefore reduced load on the heart, increasing its cardiac output. The generalised vasodilation hypothesis does not have strong grounding (19-20). It has mostly been inferred from calculating global resistance by taking arterial pressure and dividing by cardiac output and is at direct odds with microcirculatory observations. It has been demonstrated in other mathematical work that heterogeneous vessel distribution of a fixed total volume lowers the overall resistance of a vessel bed (21). Therefore microcirculatory heterogeneity may better explain the reduced systemic vascular resistance. Adequate circulatory volume is also a factor; in cases of concurrent hypovolemia patients with septic shock do not become hyperdynamic and actually develop low cardiac output with increased systemic vascular resistance (3). It can be seen from Eq. 6 that resistance of a compliant bed falls as its volume increases.

Of course, as emphasised, this is the total flow we are discussing. Assuming the ratio of forward to reverse waves is unchanged, an increase in total flow will increase the net flow (cardiac output), allowing the creation of a hyperdynamic circulation at sufficient total flow. If however wave reflections increase, this may not be the case. The Starling curve (22) demonstrates the relationship between the pressure in the right atrium (x axis) and the flow through it (y-axis) as a function of increasing cardiac fluid volume. It is curvilinear in nature. At low right atrial pressures relatively large increases in flow are associated with small increases in right atrial pressure, and at high right atrial pressures relatively small (or no) increases in flow are associated with high increases in right atrial pressure.

Therefore anything that causes either a rightward shift along the Starling curve or a rightward shift of the entire curve will increase the right atrial impedance. This will increase wave reflections. In order for cardiac output to not fall, total flow has to increase, so either the volume of the microcirculation or its flow heterogeneity has to increase. Excessive exogenous fluid administration causes a rightward shift along the curve. If the heart cannot respond to the fluid load by increasing its pumping the extra volume will not be delivered to the microcirculation. Therefore to maintain cardiac output the microvascular flow heterogeneity must increase. A failing heart, either endogenous or in response to excessive vasoconstrictor medication will result in a rightward shift of the entire curve. Again, the volume of the microvasculature cannot be increased in such a situation, and the microcirculation will become impaired if cardiac output is to be maintained. In either situation a rise in the central venous pressure without a concomitant rise in cardiac output will impair microcirculatory function. This provides a hypothetical explanation as to why high central venous pressures are an independent predictor of organ dysfunction in both sepsis and heart failure.

This is one clinical application of the hypothesis of this paper. Traditional therapies for hyperdynamic septic shock are fluid resuscitation and vasopressor medication. Fluid resuscitation, if the heart is able to deliver the administered fluid into the microcirculation, may increase the volume of the “low-volume-low-flow” microcirculatory segments, thereby increasing their flow. Vasopressors are another therapy in use, and it has never been quite clear how they should improve organ function when they constrict flow to vessel beds. In fact, vasopressors may decrease v_0_ of the “oversupplied” segments, thereby decreasing the volume required for a given flow, and this now excess volume may be shunted back to the “undersupplied” segments. However if either fluid resuscitation or vasopressor therapy is excessive and increases central venous pressure without the heart being able to pump more, then microcirculatory function is impaired, or cardiac output falls. During the development of sepsis concomitant hypovolemia and reduced microcirculatory volume may outweigh the effect of microvascular heterogeneity and total flow and cardiac output may be reduced. Administration of vasopressors may shrink the microvascular volume more than it decreases v_0_ and be insufficient to restore cardiac output. Therefore it is important to monitor both central venous pressure and indicators of cardiac output during resuscitation.

The limitations of this study is that it is theoretical and it would be difficult to develop experimental protocols to test the hypothesis. The applicability of the mathematics is related to the underlying assumptions. The non-Newtonian nature of blood may have a large effect than anticipated. There may be other physiological phenomena present that are not accounted for in the model.

## Data Availability

All data available on request

## References

1. Kislitsina ON, Rich JD, Wilcox JE, Pham DT, Churyla A, Vorovich EB, et al. Shock - Classification and Pathophysiological Principles of Therapeutics. Curr Cardiol Rev. 2019;15(2):102–113. doi: 10.2174/1573403X15666181212125024. PMID: 30543176; PMCID: PMC6520577.

2. Hernandez G, Bruhn A, Ince C. Microcirculation in sepsis: new perspectives. Curr Vasc Pharmacol. 2013 Mar 1;11(2):161–9. PMID: 23506495.

3. MacLean LD, Mulligan WG, McLean AP, Duff JH. Patterns of septic shock in man--a detailed study of 56 patients. Ann Surg. 1967 Oct;166(4):543–62. doi: 10.1097/00000658-196710000-00004. PMID: 6061542; PMCID: PMC1477449.

4. Russell JA, Rush B, Boyd J. Pathophysiology of Septic Shock. Crit Care Clin. 2018 Jan;34(1):43–61. doi: 10.1016/j.ccc.2017.08.005. PMID: 29149941.

5. Vincent JL. The International Sepsis Forum’s frontiers in sepsis: High cardiac output should be maintained in severe sepsis. Crit Care. 2003 Aug;7(4):276–8. doi: 10.1186/cc2349. Epub 2003 Jul 4. PMID: 12930548

6. Top AP, Ince C, de Meij N, van Dijk M, Tibboel D. Persistent low microcirculatory vessel density in nonsurvivors of sepsis in pediatric intensive care. Crit Care Med. 2011 Jan;39(1):8–13.

7. Rajendram R, Prowle JR. Venous congestion: are we adding insult to kidney injury in sepsis? Crit Care. 2014 Jan 27;18(1):104. doi: 10.1186/cc13709. PMID: 24467922; PMCID: PMC4055982.

8. Mullens W, Abrahams Z, Francis GS, Sokos G, Taylor DO, Starling RC, et al. Importance of venous congestion for worsening of renal function in advanced decompensated heart failure. J Am Coll Cardiol 2009, 53: 589-596. 10.1016/j.jacc.2008.05.068

9. O’Rourke MF. Wave travel and reflection in the arterial system. J Hypertens Suppl. 1999 Dec;17(5):S45–7. PMID: 10706326.

10. Naishadham K. Transmission Lines. In: Chen W, editors. The Electrical Engineering Handbook. Academic Press; 2005. p 525–537. ISBN 9780121709600.

11. Secomb TW. Mechanics of blood flow in the microcirculation. Symp Soc Exp Biol. 1995;49:305–21. PMID: 8571232.

12. Guyton AC, Lindsey AW, Abernathy B, Richardson T. Venous return at various right atrial pressures and the normal venous return curve. Am J Physiol. 1957 Jun;189(3):609–15. doi: 10.1152/ajplegacy.1957.189.3.609. PMID: 13458395.

13. Magder S, Famulari G, Gariepy B. Periodicity, time constants of drainage, and the mechanical determinants of peak cardiac output during exercise. J Appl Physiol (1985). 2019 Dec 1;127(6):1611–1619. doi: 10.1152/japplphysiol.00688.2018. Epub 2019 Aug 15. PMID: 31414960.

14. Pedley, T., Luo, X. Modelling Flow and Oscillations in Collapsible Tubes. Theoret. Comput. Fluid Dynamics 10, 277–294 (1998). 10.1007/s001620050064

15. Kozlovsky P, Zaretsky U, Jaffa AJ, Elad D. General tube law for collapsible thin and thick-wall tubes. J Biomech. 2014 Jul 18;47(10):2378–84. doi: 10.1016/j.jbiomech.2014.04.033. Epub 2014 Apr 26. PMID: 24837222.

16. Secomb TW. Mechanics of blood flow in the microcirculation. Symp Soc Exp Biol. 1995;49:305–21. PMID: 8571232.

17. Alotta G, Bologna E, Failla G, Zingales M. A Fractional Approach to Non-Newtonian Blood Rheology in Capillary Vessels. J Peridyn Nonlocal Model. 2019;1:88–96. doi:10.1007/s42102-019-00007-9

18. Kennelly AE. Time Constants for Engineering Purposes in Simple Exponential Transient Phenomena. Proc Natl Acad Sci U S A. 1924 Nov;10(11):467–73. doi: 10.1073/pnas.10.11.467. PMID: 16586952; PMCID: PMC1085755.

19. Tucker JJ, Wilson MA, Wead WB, Garrison RN. Microvascular endothelial cell control of peripheral vascular resistance during sepsis. Arch Surg. 1998 Dec;133(12):1335–42. doi: 10.1001/archsurg.133.12.1335. PMID: 9865652.

20. Tyml K, Yu J, McCormack DG. Capillary and arteriolar responses to local vasodilators are impaired in a rat model of sepsis. J Appl Physiol (1985). 1998 Mar;84(3):837–44. doi: 10.1152/jappl.1998.84.3.837. PMID: 9480941.

21. Rosenberg E. On deriving Murray’s law from constrained minimization of flow resistance. J Theor Biol. 2021 Mar 7;512:110563. doi: 10.1016/j.jtbi.2020.110563. Epub 2020 Dec 24. PMID: 33359240.

22. Berlin DA, Bakker J. Starling curves and central venous pressure. Crit Care. 2015 Feb 16;19(1):55. doi: 10.1186/s13054-015-0776-1. PMID: 25880040; PMCID: PMC4329649.

